# Nutritional Risk Screening and Clinical Outcomes in a Tertiary University Hospital: A Real-World Retrospective Cohort Study

**DOI:** 10.64898/2026.09.19.26363474

**Authors:** Marie Gerwek, Silvia Walther, Ulrike Holdgrün, Alicia Nierling, Lars Selig, Michael Stumvoll, Toralf Kirsten, Haiko Schlögl

## Abstract

**Background & objectives:** Malnutrition risk is common among hospitalized patients and is associated with adverse clinical outcomes. However, implementation of routine nutritional screening remains heterogeneous. This study assessed the distribution and implementation of Nutritional Risk Screening 2002 (NRS-2002) and examined associations between malnutrition risk and clinical outcomes in a tertiary university hospital.

**Methods:** We conducted a retrospective cohort study using routinely collected electronic health record data from adult hospitalizations at the University of Leipzig Medical Center, Germany, between 2013 and 2023. The implementation of the NRS-2002 in 2017 enabled comparison of a main cohort with potential NRS assessment (2017–2023) and a pre-implementation comparison cohort (2013–2016). In the main cohort, hospitalizations were categorized according to screening results as no elevated malnutrition risk, NRS ≥ 3 points, or positive initial screening without a documented final NRS assessment. Clinical outcomes included 30-day hospital readmission, intensive care unit (ICU) admission, mechanical ventilation, in-hospital mortality, and length of hospital stay (LOS). Associations were assessed using unadjusted and age- and sex-adjusted logistic regression. Length of hospital stay was analyzed using Gamma regression. Time to readmission was additionally assessed using Kaplan–Meier and Cox regression.

**Results:** After data preprocessing, 178,607 hospitalizations were included in the main cohort and 52,044 in the comparison cohort. In the main cohort, 5,460 hospitalizations (3.1%) had an NRS ≥3 points, while 21,842 (12.2%) had a positive initial screening without a documented final assessment. Patients with NRS ≥3 points had higher rates of 30-day readmission, ICU admission, and in-hospital mortality and longer LOS than patients without elevated malnutrition risk. After adjustment for age and sex, NRS ≥3 points remained associated with higher odds of 30-day readmission (OR 1.57, 95% CI 1.39–1.77), ICU admission (OR 1.85, 95% CI 1.63–2.11), in-hospital mortality (OR 3.62, 95% CI 2.69–4.99), and mechanical ventilation (OR 2.07, 95% CI 1.08–4.38). Hospitalizations with a positive initial screening but without a documented final assessment also showed increased odds of several adverse outcomes, including 30-day readmission and in-hospital mortality. The proportion of positive initial screenings and completion of the final NRS assessment varied substantially across clinical departments.

**Conclusion:** Elevated malnutrition risk was associated with adverse clinical outcomes in this large real-world cohort. The substantial proportion of positive initial screenings without subsequent final assessment highlights important challenges in implementing a complete nutritional screening pathway in routine hospital care.

## Introduction

Disease-related malnutrition remains a critical challenge in acute care and is associated with increased in-hospital mortality, prolonged length of stay (LOS), and substantial healthcare expenditures [1, 2]. European registry data and multi-center studies have estimated that approximately 30% of hospitalized adults are at risk of malnutrition, with disproportionately high rates among older adults and patients with cancer [3, 4, 5, 6, 7]. To identify patients at malnutrition risk at an early stage, international guidelines from the European Society for Clinical Nutrition and Metabolism (ESPEN) recommend systematic nutritional screening followed by appropriate nutritional assessment and intervention. The Nutritional Risk Screening 2002 (NRS-2002) is a widely validated screening tool for identifying malnutrition risk in hospitalized patients [8, 9]. Using a two-step approach, it evaluates nutritional status based on BMI, recent weight loss, and reduced dietary intake alongside disease severity, with an additional age adjustment for patients aged 70 years or older. A cumulative score of 3 or higher indicates malnutrition risk and identifies patients who may benefit from further nutritional assessment and intervention.

Earlier evidence from the German Hospital Malnutrition Study identified higher age, malignant disease, and admission to geriatric or oncology departments as characteristics associated with hospital malnutrition in Germany. Furthermore, malnutrition was associated with a 43% longer hospital stay, emphasizing its considerable clinical and healthcare burden [3]. More recently, the EFFORT trial provided evidence that individualized, protocol-driven nutritional therapy for hospitalized patients at malnutrition risk can reduce adverse clinical outcomes, including severe complications and mortality [10].

Despite evidence supporting nutritional screening and intervention, routine implementation of nutritional screening in clinical practice remains fragmented across European healthcare systems. In Germany, structural hurdles, including historical gaps in reimbursement for malnutrition diagnoses, have been reported as barriers to widespread adoption. While a nationwide survey indicated that 72% of German hospitals conduct some form of nutritional screening, only 40% reported systematic screening across all departments [11]. Similarly, a national survey of patients with cancer found that only 59% of participating institutions reported nutritional therapy as an integral part of oncological treatment and 65.1% reported routinely conducting nutritional screening [12]. These findings highlight a persistent gap between guideline recommendations and their implementation in routine clinical practice. Furthermore, existing German epidemiological data are largely derived from selected geriatric, oncological, or surgical populations, limiting the assessment of malnutrition risk across broad inpatient populations.

To address these knowledge gaps, we conducted a hospital-wide real-world evaluation of routine nutritional screening using routinely collected patient data from a large tertiary university hospital in Germany. We assessed the prevalence and distribution of malnutrition risk, evaluated implementation of the intended NRS-2002 screening pathway across clinical departments, and examined associations between malnutrition risk and major clinical outcomes, including in-hospital mortality, length of stay, intensive care unit (ICU) admission, mechanical ventilation, and 30-day readmission. We hypothesized that elevated malnutrition risk would be associated with adverse clinical outcomes and that implementation of the screening pathway would vary substantially across clinical departments.

## Material & Methods

### Study Design and Objectives

This retrospective cohort study utilized routinely collected electronic health record (EHR) data from the University of Leipzig Medical Center (ULMC), a tertiary care academic hospital in Germany. The primary objective was to determine the hospital-wide prevalence and distribution of malnutrition risk among adult inpatients and to evaluate the implementation of the routine NRS-2002 screening pathway across clinical departments. The secondary objective was to examine associations between malnutrition risk and clinical outcomes, including in-hospital mortality, 30-day readmission, intensive care unit (ICU) admission, mechanical ventilation, and length of hospital stay (LOS). The study protocol was approved by the Ethics Committee of the University of Leipzig (Ref. No. 12/24-ek) and conducted in accordance with the Declaration of Helsinki.

### Study Population and Eligibility Criteria

Data were extracted for all adult patients (≥ 18 years) admitted to ULMC between January 1, 2013, and December 31, 2023. The initial dataset comprised 556,789 hospitalizations among 276,223 individual patients (mean age at admission: 57.7±19.3 years; 49% female). The study size was determined by the availability of eligible hospitalizations within the predefined study period and data sources. Unique patient identifiers enabled longitudinal linkage across multiple hospitalizations. The unit of analysis was the hospitalization.

### Nutritional Risk Assessment

Malnutrition risk was assessed using the Nutritional Risk Screening 2002 (NRS-2002), which was implemented at ULMC in 2017. The screening follows a two-stage workflow consisting of an initial screening followed, if indicated, by a final NRS-2002 assessment. The initial screening is conducted at admission by administrative personnel using four binary (yes/no) items assessing low BMI, recent weight loss, reduced food intake, and severe illness. A positive initial screening triggered an automated notification to the clinical nutrition team. The final NRS-2002 assessment was conducted by trained clinical nutrition specialists for patients flagged by the initial screening. The final NRS-2002 score combines impaired nutritional status (0–3 points), disease severity (0–3 points), and an age adjustment (+1 point for age ≥ 70 years). For descriptive analyses, final NRS-2002 scores were categorized as no/low risk (0–2 points), moderate risk (3–4 points), or high risk (≥ 5 points) [13]. For outcome analyses, hospitalizations in the main cohort were grouped into three categories: (1) no elevated malnutrition risk, comprising a negative initial screening or a final NRS-2002 score of 0–2; (2) elevated malnutrition risk, defined as a final NRS-2002 score ≥ 3; and (3) positive initial screening only, defined as a positive initial screening without a documented final NRS-2002 assessment. The no-elevated-risk group served as the reference category. Because final NRS-2002 assessments were not completed for all hospitalizations with a positive initial screening, the observed prevalence of elevated malnutrition risk may underestimate the prevalence in the screened population. Therefore, an extrapolated prevalence was calculated by applying the observed proportion of NRS ≥ 3 points among completed assessments to hospitalizations with a positive initial screening but no documented final assessment. In the comparison cohort, the corresponding pre-implementation screening status was based on the routinely documented pre-screening procedure used before implementation of the NRS-2002.

### Data Extraction, Management, and Preprocessing

Data integration was conducted by the Data Integration Center, a dedicated unit of the ULMC, which extracted the requested data from the enterprise hospital information system [14] and provided them in raw format for analysis. All data processing was performed within the premises of the ULMC using dedicated computing infrastructure. Data processing and standardization were performed using R software (v4.5.1; [15]). The dataset included demographic and administrative information, diagnoses and procedures, laboratory measurements, and nutritional screening data. Unstructured NRS documentation was parsed and transformed into structured tabular data, and extracted NRS scores were algorithmically checked for internal consistency. Each hospitalization represented a complete inpatient stay from admission to discharge. Patients were assigned a unique identifier at their first admission, enabling longitudinal linkage across multiple hospitalizations. Automated data quality checks were performed to assess data completeness, plausible value ranges, and distributional characteristics.

To define the study population and ensure data quality, hospitalizations were excluded in several steps. Hospitalizations with documented ascites or a length of stay (LOS) of less than 1 day or more than 180 days were excluded. Laboratory values recorded within 14 days prior to bariatric procedures or during active dialysis treatment were excluded from the respective laboratory-based analyses. Hospitalizations that were not assigned to an eligible adult discharge unit, obstetrics, intensive care unit, emergency department, or day clinic were subsequently excluded. Additional records with inconsistent or implausible data were removed according to predefined variable-specific criteria. These criteria included fixed plausible ranges for variables such as BMI, LOS, and readmission-related measures. For laboratory variables, extreme values exceeding three standard deviations from the respective variable mean were considered outliers and excluded. For each hospitalization, the last available measurement during the relevant hospitalization was retained for laboratory variables.

Hospitalizations with missing key variables required for the respective analyses, including age, sex, admission or discharge date, primary diagnosis, initial screening status, ICU admission, or in-hospital mortality, were excluded.

The Charlson Comorbidity Index was calculated using the Comorbidity R package [16]. Primary diagnoses were grouped according to selected chapters of the International Classification of Diseases, 10th Revision (ICD-10). These variables were used to characterize the clinical composition of the study population. Each hospitalization was assigned to a medical specialty according to the final discharge unit. Discharge units were used to describe the distribution of nutritional screening and screening completion across clinical departments.

### Statistical Analysis

Continuous variables were assessed for distributional characteristics and expressed as mean ± standard deviation (SD) or median with interquartile range (IQR), as appropriate. Categorical variables are presented as absolute counts and percentages. Descriptive statistics were calculated to characterize the study population. The distribution of malnutrition risk was described across age groups, and across discharge units.

Associations between malnutrition risk categories and binary clinical outcomes, including 30-day readmission, mechanical ventilation, in-hospital mortality, and ICU admission, were evaluated using logistic regression models. Odds ratios (ORs) with 95% confidence intervals (CIs) were reported. Two models were fitted for each outcome: an unadjusted model and a model adjusted for age and sex. Length of hospital stay was analyzed as a continuous outcome using Gamma regression with a log link. Regression analyses were performed using complete hospitalizations for the variables included in each respective model.

Time to 30-day readmission was additionally evaluated using Kaplan–Meier survival analysis and Cox proportional hazards regression. Statistical significance was defined as a two-sided *P* < 0.05.

## Results

### Cohort Definitions

The study population was divided into a main cohort comprising hospitalizations from 2017 to 2023, after implementation of the NRS-2002 at ULMC, and a comparison cohort comprising hospitalizations from 2013 to 2016, before implementation of the NRS-2002 (Figure 1). The cohorts were separated based on admission date. There were 569 hospitalizations with overlapping stays, which were assigned to the comparison cohort. After data preprocessing, 178,607 hospitalizations remained in the main cohort (mean age: 59.1 ± 19.2 years; 48.9% female), while 52,044 remained in the comparison cohort (mean age: 57.4 ± 18.3 years; 48.4% female).

**Figure 1:**
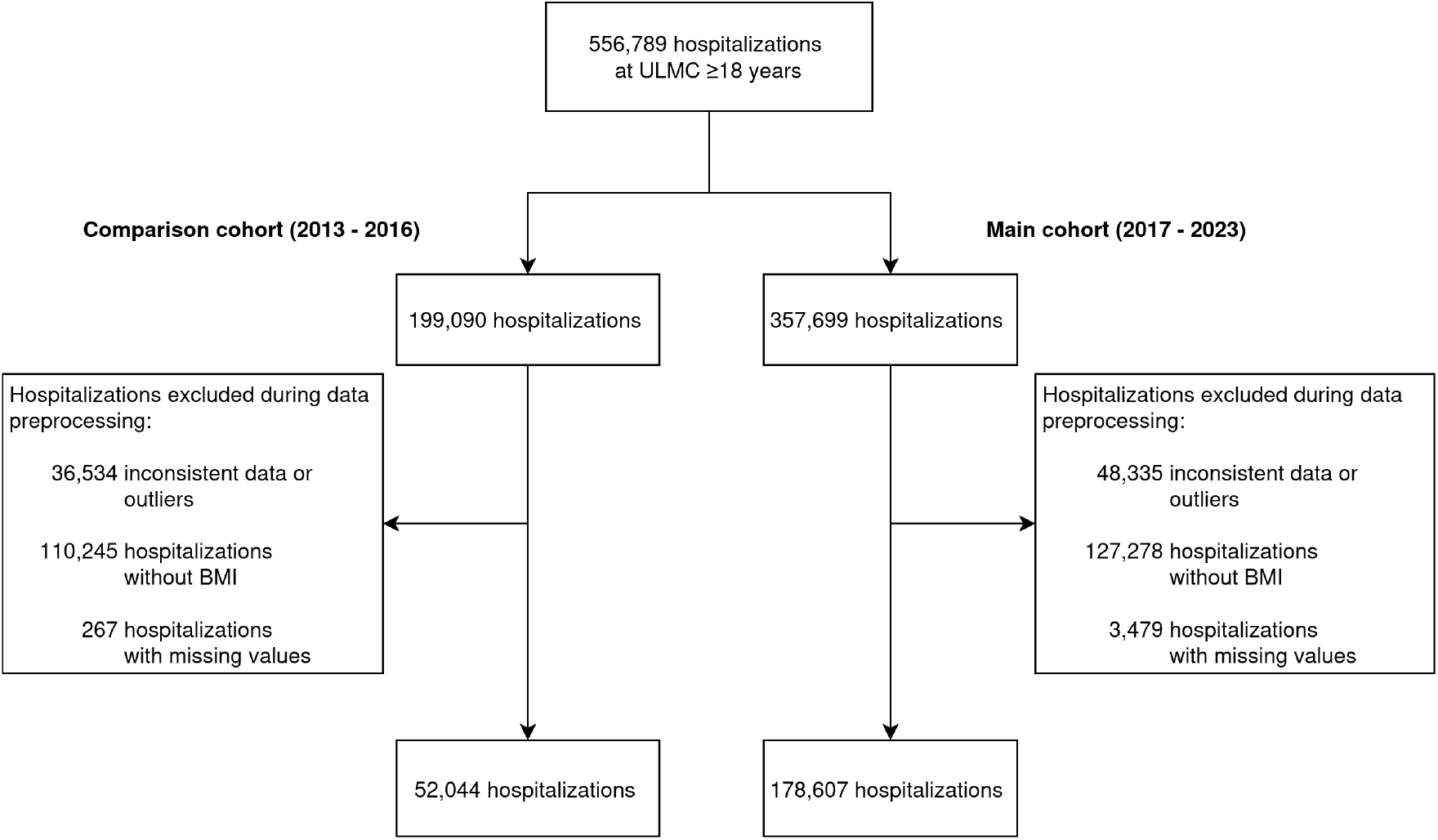
Flowchart of cohort selection and data preprocessing. All hospitalized adult hospitalizations (≥ 18 years) at the University Leipzig Medical Center (ULMC) between January 2013 and December 2023 were initially identified. The study population was divided into a comparison cohort (2013–2016) and a main cohort (2017–2023) based on the implementation of Nutritional Risk Screening 2002 at the ULMC. Subsequent data preprocessing included removal of implausible values and outliers, exclusion of hospitalizations without BMI, and exclusion of hospitalizations with missing key variables.

In the main cohort, hospitalizations were categorized according to the initial and final screening results. Most hospitalizations had a negative initial screening (n=148,305; 83.0%). A final NRS-2002 assessment documented no elevated malnutrition risk (NRS ≤2 points) in 3,000 hospitalizations (1.7%) and elevated malnutrition risk (NRS ≥3 points) in 5,460 (3.1%). A further 21,842 hospitalizations (12.2%) had a positive initial screening but no documented final NRS-2002 assessment. In the comparison cohort, 8,073 hospitalizations (15.5%) had a positive and 43,971 (84.5%) a negative initial screening.

### Baseline Characteristics

Baseline characteristics differed between the nutritional screening groups (Table 1). In the main cohort, hospitalizations with elevated malnutrition risk (NRS ≥3 points) involved older patients than those with NRS ≤2 points (mean age: 64.3 ±15.6 vs. 55.4 ±17.1 years) and had a lower proportion of females (45.6% vs. 50.9%). Among hospitalizations with an initial screening only, females were more frequently represented in the positive screening group than in the negative screening group, accounting for 55.5% of hospitalizations in the main cohort and 54.1% in the comparison cohort. In both cohorts, hospitalizations with a negative initial screening involved older patients than those with a positive initial screening (main cohort: 59.4 ±19.1 vs. 56.6 ±20.6 years; comparison cohort: 58.0 ±18.1 vs. 54.3 ±19.2 years).

**Table 1.** Baseline characteristics of the study population stratified by malnutrition risk status and cohort. The main cohort includes hospitalizations categorized according to the availability and result of the final NRS-2002 assessment and the initial screening result. Hospitalizations with a documented final assessment were categorized as NRS ≤2 or NRS ≥3 points, while hospitalizations without a documented final assessment were categorized according to the initial screening result. The comparison cohort comprises hospitalizations before implementation of the NRS-2002 and is similarly stratified by pre-implementation screening status. Comorbidities were defined according to routine clinical documentation, and overall comorbidity burden was assessed using the Charlson Comorbidity Index. NRS: Nutritional Risk Screening 2002.

|  | Main cohort (2017–2023) |  |  |  | Comparison cohort (2013–2016) |  |
| --- | --- | --- | --- | --- | --- | --- |
|  | NRS ≤2 points<br>n=3,000 | NRS ≥3 points<br>n=5,460 | Only positive<br>initial screening<br>n=21,842 | Negative<br>initial screening<br>n=148,305 | Positive<br>initial screening<br>n=8,073 | Negative<br>initial screening<br>n=43,971 |
| *Sociodemographics* |  |  |  |  |  |  |
| Age at admission (years), mean (SD) | 55.4 (17.1) | 64.3 (15.6) | 56.6 (20.6) | 59.4 (19.1) | 54.3 (19.2) | 58.0 (18.1) |
| Sex, % (total) |  |  |  |  |  |  |
| Female | 50.9% (1,526) | 45.6% (2,491) | 55.5% (12,125) | 48.0% (71,138) | 54.1% (4,370) | 47.4% (20,837) |
| Male | 49.1% (1,474) | 54.4% (2,969) | 44.5% (9,717) | 52.0% (77,167) | 45.9% (3,703) | 52.6% (23,134) |
| *Nutritional assessment, mean (SD)* |  |  |  |  |  |  |
| Body-mass index (kg/m<sup>2</sup>) | 25.8 (10.0) | 22.4 (5.9) | 21.8 (4.8) | 27.7 (5.5) | 21.8 (5.3) | 27.5 (5.6) |
| Serum albumin (g/l) | 38.0 (6.9) | 32.9 (7.1) | 36.7 (7.2) | 39.7 (6.5) | 37.2 (7.2) | 40.1 (6.3) |
| Serum phosphate (mmol/l) | 1.1 (0.3) | 1.1 (0.3) | 1.1 (0.3) | 1.1 (0.3) | 1.1 (0.3) | 1.1 (0.3) |
| *Comorbidities, % (total)* |  |  |  |  |  |  |
| Coronary heart disease | 1.9% (57) | 3.3% (181) | 2.9% (631) | 4.2% (6,191) | 1.0% (84) | 1.8% (771) |
| Congestive heart failure | 6.4% (192) | 10.9% (596) | 8.2% (1,787) | 9.7% (14,375) | 5.5% (447) | 5.8% (2,561) |
| Peripheral arterial disease | 4.8% (143) | 7.5% (409) | 5.1% (1,111) | 5.8% (8,550) | 4.3% (344) | 5.3% (2,321) |
| Cerebrovascular disease | 2.6% (77) | 5.9% (322) | 6.0% (1,301) | 6.2% (9,158) | 4.0% (320) | 4.5% (1,958) |
| Dementia | 1.1% (32) | 3.8% (206) | 4.2% (928) | 3.0% (4,482) | 2.0% (159) | 1.8% (788) |
| Chronic obstructive pulmonary disease | 9.1% (272) | 10.4% (570) | 7.8% (1,698) | 7.2% (10,675) | 6.8% (553) | 5.6% (2,468) |
| Diabetes | 17.2% (515) | 19.9% (1,085) | 15.0% (3,279) | 20.2% (29,891) | 12.7% (1,026) | 17.3% (7,595) |
| Chronic kidney disease | 11.3% (339) | 16.3% (890) | 13.4% (2,937) | 14.7% (21,751) | 11.3% (916) | 12.4% (5,442) |
| Malignant disease | 33.9% (1,018) | 50.1% (2,736) | 26.7% (5,834) | 14.3% (21,259) | 32.3% (2,606) | 19.5% (8,587) |
| Charlson comorbidity index, mean (SD) | 2.3 (2.6) | 3.4 (2.8) | 2.2 (2.5) | 1.7 (2.1) | 2.3 (2.6) | 1.7 (2.2) |
| *Main diagnosis, % (total)* |  |  |  |  |  |  |
| Blood diseases | 4.0% (94) | 3.2% (143) | 4.2% (668) | 4.9% (4,170) | 5.3% (309) | 5.3% (1,361) |
| Circulatory system | 7.4% (173) | 7.1% (317) | 12.4% (1,943) | 30.0% (25,767) | 7.0% (409) | 19.2% (4,933) |
| Digestive diseases | 22.2% (523) | 17.4% (774) | 13.7% (2,149) | 10.4% (8,941) | 16.5% (959) | 11.9% (3,069) |
| Endocrine diseases | 12.8% (301) | 2.1% (93) | 3.7% (580) | 4.8% (4,154) | 4.0% (234) | 7.1% (1,837) |
| Infectious diseases | 2.4% (57) | 3.8% (171) | 4.6% (724) | 4.1% (3,549) | 3.2% (188) | 2.9% (745) |
| Mental disorders | 0.6% (15) | 0.5% (24) | 9.9% (1,561) | 6.4% (5,464) | 7.5% (435) | 5.9% (1,515) |
| Neoplasms | 42.5% (1,000) | 57.1% (2,539) | 38.6% (6,073) | 24.4% (20,964) | 47.6% (2,765) | 35.9% (9,214) |
| Nervous system | 2.2% (51) | 2.3% (101) | 5.0% (793) | 7.3% (6,280) | 4.9% (286) | 6.3% (1,623) |
| Respiratory diseases | 5.9% (139) | 6.4% (284) | 7.8% (1,227) | 7.6% (6,521) | 3.9% (228) | 5.5% (1,401) |

Markers of nutritional status also differed between screening groups. In the main cohort, hospitalizations with NRS ≥3 points had lower BMI and serum albumin levels than those with NRS ≤2 points (22.4 vs. 25.8 kg/m^2^ and 32.9 vs. 38.0 g/L, respectively). Among hospitalizations without a documented final NRS-2002 assessment, those with a positive initial screening also had lower BMI and serum albumin levels than those with a negative initial screening. This pattern was observed in both the main cohort (BMI: 21.8 vs. 27.7 kg/m^2^; serum albumin: 36.7 vs. 39.7 g/L) and the comparison cohort (BMI: 21.8 vs. 27.5 kg/m^2^; serum albumin: 37.2 vs. 40.1 g/L).

Hospitalizations with NRS ≥3 points had a higher comorbidity burden, with a mean Charlson Comorbidity Index of 3.4. Nearly half of these hospitalizations involved patients with a malignant disease (50.1%), and neoplasms accounted for 57.1% of primary diagnoses. Other frequently observed comorbidities included diabetes (19.9%) and chronic kidney disease (16.3%). Detailed baseline characteristics of the study population are presented in Table 1.

### NRS-2002 Implementation

The proportion of positive initial screenings varied substantially across clinical departments. The highest rates were observed in palliative care (57.7%), oncology (48.2%), psychology (45.9%), infectious diseases (44.1%), and radiotherapy (42.8%). Overall, 21,842 hospitalizations had a positive initial screening but no documented final NRS-2002 assessment.

Among hospitalizations with a positive initial screening, completion of the final NRS-2002 assessment also varied substantially across departments. The highest completion rates were observed in visceral surgery (37.0%) and radiotherapy (35.4%), whereas completion was lowest in psychiatry (0.3%) and psychology (1.3%) (Supplementary Figure 4).

### Distribution of Malnutrition Risk

Among hospitalizations with a documented final NRS-2002 assessment, the prevalence of elevated malnutrition risk increased with age. The highest prevalence was observed in patients aged 70–79 years, with a slight decrease among patients aged 80 years and older. Across age groups, high malnutrition risk (NRS ≥5) accounted for approximately 10–15% of completed assessments, while the combined prevalence of moderate and high malnutrition risk increased substantially with age, reaching up to 80% among patients aged ≥70 years (Figure 2).

**Figure 2:**
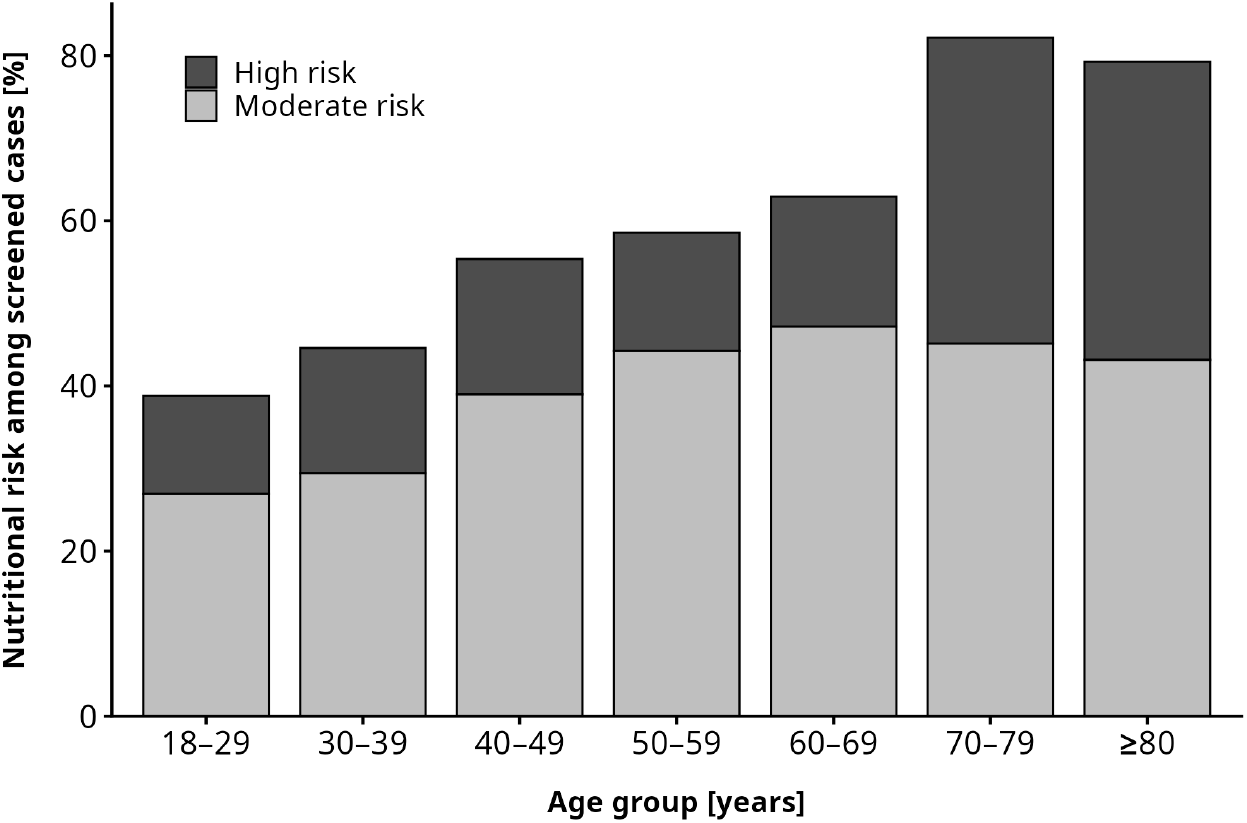
Prevalence of malnutrition risk stratified by age group. Bars represent the proportion of hospitalizations at moderate malnutrition risk (NRS 3–4 points) and high malnutrition risk (NRS≥5 points) among all hospitalizations that received a final screening. NRS: Nutritional Risk Screening 2002.

The extrapolated prevalence of elevated malnutrition risk varied considerably across discharge units. The highest proportions were estimated in oncology (53.6%, 95% CI 51.9–55.1) and palliative care (52.6%, 95% CI 49.4–54.9), followed by radiotherapy (35.2%, 95% CI 33.3–36.9) and gastroenterology (28.0%, 95% CI 26.4–29.5) (Figure 3). These findings indicate a concentration of malnutrition risk in patient populations with severe and chronic underlying conditions.

**Figure 3:**
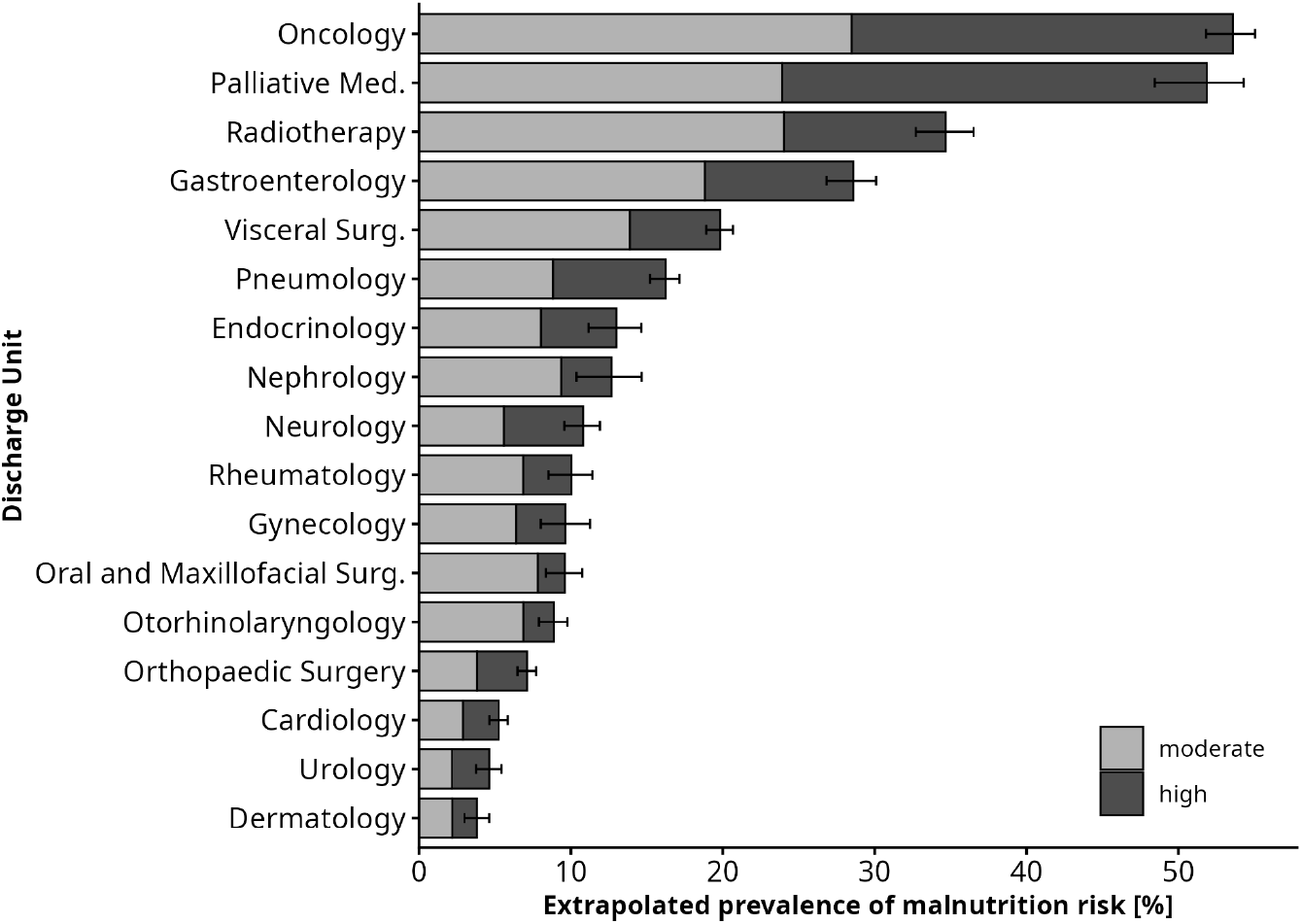
Extrapolated prevalence of malnutrition risk by discharge unit. Bars show the estimated prevalence of moderate (NRS 3–4 points) and high malnutrition risk (NRS ≥5 points) after extrapolating the observed distribution among hospitalizations with a positive initial screening and documented final NRS-2002 assessment to hospitalizations with a positive initial screening but no documented final assessment. Error bars represent 95% confidence intervals for the total estimated prevalence. Units with fewer than 100 hospitalizations or fewer than 50 hospitalizations with both a positive initial screening and completed final assessment were excluded. NRS: Nutritional Risk Screening 2002.

### Clinical Outcomes

Clinical outcomes differed across the nutritional screening groups (Table 2). In the main cohort, hospitalizations with NRS ≥3 points generally showed higher frequencies of adverse outcomes than those with NRS ≤2 points or a negative initial screening. Hospitalizations with a positive initial screening but without a documented final NRS-2002 assessment showed intermediate outcome frequencies for several outcomes. In the comparison cohort, hospitalizations with a positive initial screening also showed higher frequencies of 30-day readmission and in-hospital mortality than those with a negative initial screening, whereas ICU admission and mechanical ventilation occurred at similar frequencies between the two screening groups.

**Table 2.**
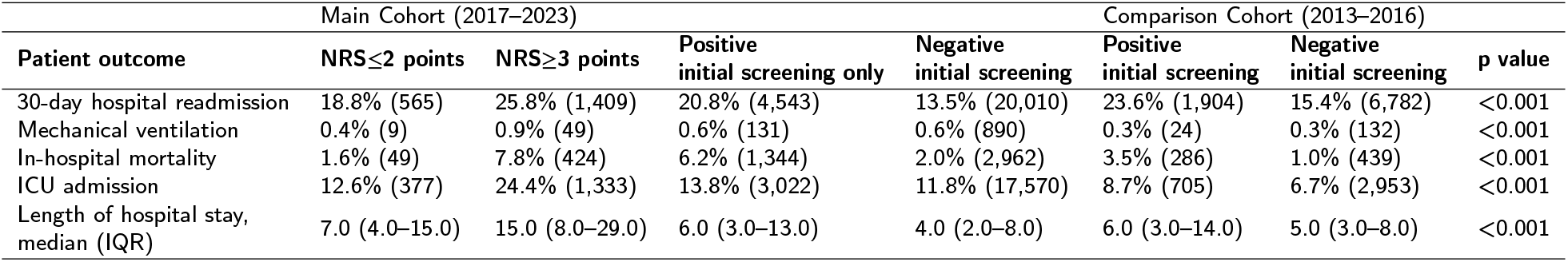
Clinical outcomes stratified by nutritional screening group. Hospitalizations were categorized according to the availability and result of the final NRS-2002 assessment and the initial screening result during the main and comparison periods. Binary outcomes are presented as percentages with absolute counts, and continuous variables as median (interquartile range). P values represent global comparisons across all six groups and were calculated using chi-squared tests for categorical variables and the Kruskal–Wallis test for continuous variables. NRS: Nutritional Risk Screening 2002.

In the main cohort, 30-day hospital readmission occurred in 25.8% of hospitalizations with NRS ≥3 points, compared with 18.8% among those with NRS ≤2 points and 13.5% among those with a negative initial screening. ICU admission occurred in 24.4%, 12.6%, and 11.8% of these groups, respectively. In-hospital mortality was 7.8% among hospitalizations with NRS ≥3 points, compared with 1.6% among those with NRS ≤2 points and 2.0% among those with a negative initial screening. Mechanical ventilation occurred infrequently across all groups, but was more frequent among hospitalizations with NRS ≥3 points (0.9%) than among those with NRS ≤2 points (0.4%) or a negative initial screening (0.6%). In the comparison cohort, 30-day readmission and in-hospital mortality were more frequent among hospitalizations with a positive initial screening than among those with a negative initial screening (23.6% vs. 15.4% and 3.5% vs. 1.0%, respectively), whereas ICU admission (8.7% vs. 6.7%) and mechanical ventilation showed no differences.

Length of hospital stay also differed substantially across final NRS-2002 categories. Patients with NRS ≤2 points had a median LOS of 7 days (IQR 4–15), compared with 14 days (IQR 7–28) among patients with NRS 3–4 points and 15 days (IQR 8–30) among those with NRS ≥5 points (Kruskal–Wallis *p* < 0.001) (Figure 4).

**Figure 4:**
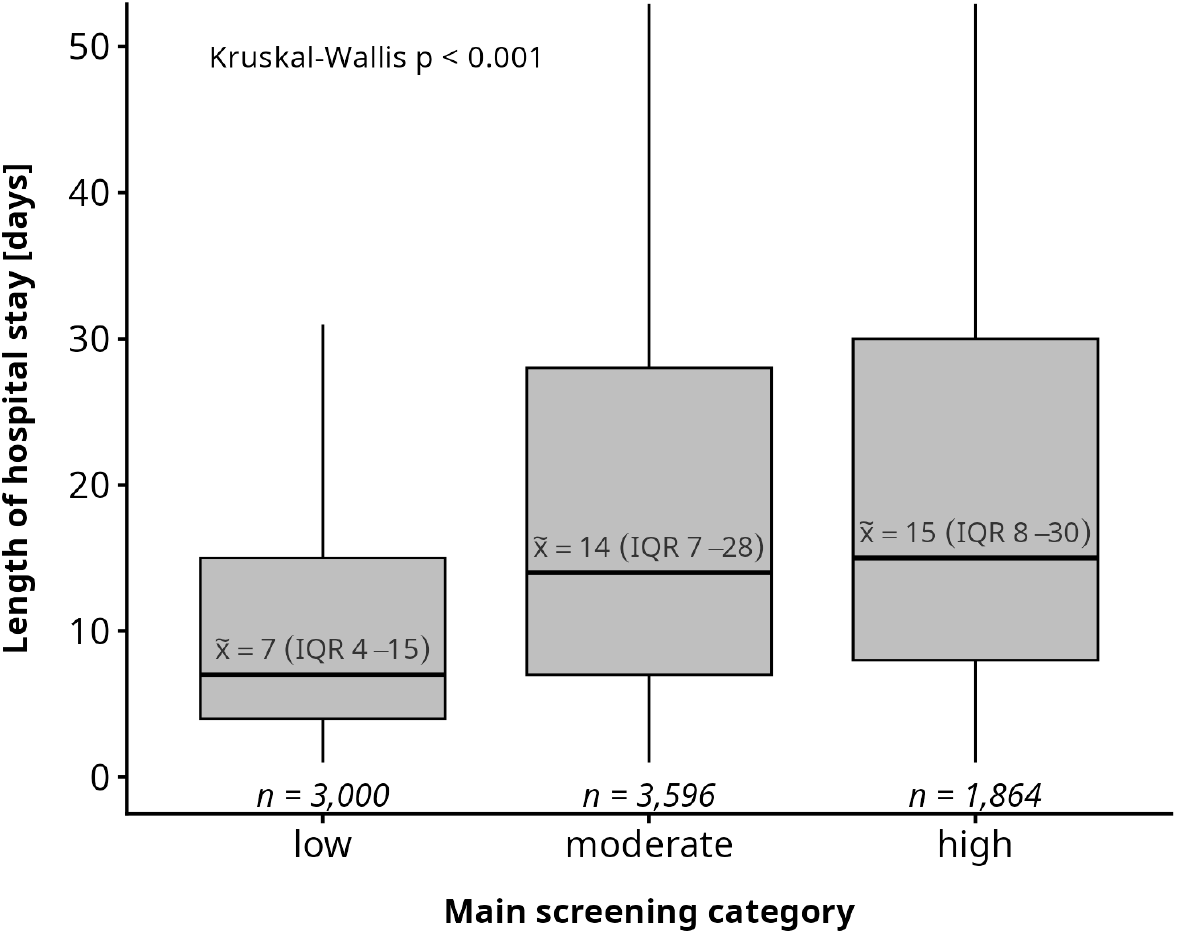
Length of hospital stay in days stratified by final screening category based on NRS-2002. Patients were categorized into low (NRS ≤ 2 points), moderate (NRS 3–4 points), and high (NRS ≥ 5 points) malnutrition risk according to the final screening. Median values and interquartile ranges are annotated within each group, and sample sizes are shown below. Differences between groups were assessed using the Kruskal–Wallis test. The y-axis is truncated at the 95th percentile. NRS: Nutritional Risk Screening 2002.

Kaplan–Meier analysis demonstrated differences in time to 30-day hospital readmission between malnutrition risk groups (Figure 5). Compared with the reference group without elevated malnutrition risk, the unadjusted Cox model showed higher hazards of 30-day readmission among hospitalizations with NRS ≥3 points (HR 1.73, 95% CI 1.70– 1.75; *p* < 0.001) and those with a positive initial screening but no documented final NRS-2002 assessment (HR 1.46, 95% CI 1.44–1.47; *p* < 0.001).

**Figure 5:**
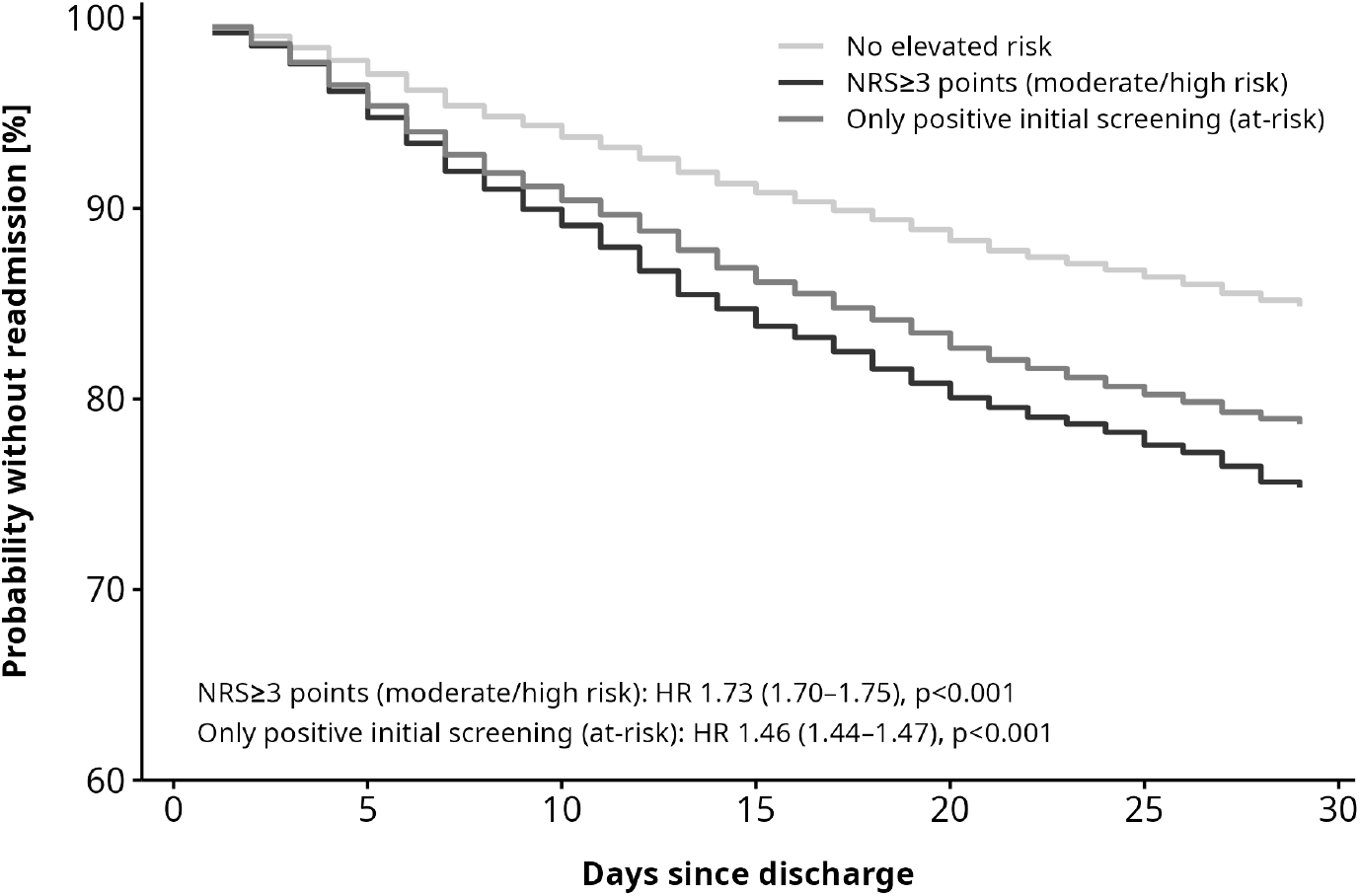
Kaplan–Meier curves for time to hospital readmission within 30 days after discharge, stratified by malnutrition risk group. Patients without a readmission during the 30-day follow-up period were censored at the end of follow-up. The reference group comprised hospitalizations without elevated malnutrition risk (negative initial screening or NRS ≤2 points). Hazard ratios (HRs) with 95% confidence intervals were estimated using an unadjusted Cox proportional hazards model. NRS: Nutritional Risk Screening 2002.

Logistic regression analyses showed associations between malnutrition risk and several clinical outcomes (Table 3). In the age- and sex-adjusted models, hospitalizations with NRS ≥3 points had higher odds of 30-day readmission (OR 1.57, 95% CI 1.39–1.77), ICU admission (OR 1.85, 95% CI 1.63–2.11), and in-hospital mortality (OR 3.62, 95% CI 2.69–4.99) compared with the reference group. NRS ≥3 points was also associated with higher odds of mechanical ventilation (OR 2.07, 95% CI 1.08–4.38, *P* = 0.039). For hospitalizations with a positive initial screening only, adjusted odds were increased for 30-day readmission (OR 1.21, 95% CI 1.09–1.35) and in-hospital mortality (OR 3.46, 95% CI 2.60–4.71), but not for ICU admission (OR 1.06, 95% CI 0.94–1.20, *P* = 0.340) or mechanical ventilation (OR 1.48, 95% CI 0.82–3.03, *P* = 0.234).

**Table 3.** Association between malnutrition risk and clinical outcomes in main cohort. Odds ratios (ORs) with 95% confidence intervals (CIs) are presented for 30-day hospital readmission, in-hospital mortality, mechanical ventilation, and ICU admission. Estimates are shown for hospitalizations with NRS ≥3 points and for hospitalizations with a positive initial screening without a documented final NRS-2002 assessment, each compared with the reference group without elevated malnutrition risk. Unadjusted and age- and sex-adjusted logistic regression models are reported. CI: confidence interval; ICU: intensive care unit; NRS: Nutritional Risk Screening 2002; OR: odds ratio.

| Outcome | Unadjusted OR | Adjusted OR |
| --- | --- | --- |
|  | (95% CI), p value | (95% CI), p value (age/sex) |
| *30-day readmission* |  |  |
| NRS $\geq 3$ points | 1.555 (1.382–1.751), $p < 0.001$ | 1.569 (1.393–1.769), $p < 0.001$ |
| Only positive initial screening | 1.181 (1.064–1.314), $p = 0.002$ | 1.214 (1.093–1.351), $p < 0.001$ |
| *In-hospital mortality* |  |  |
| NRS $\geq 3$ points | 4.802 (3.574–6.605), $p < 0.001$ | 3.624 (2.692–4.994), $p < 0.001$ |
| Only positive initial screening | 3.744 (2.824–5.093), $p < 0.001$ | 3.456 (2.603–4.708), $p < 0.001$ |
| *Mechanical ventilation* |  |  |
| NRS $\geq 3$ points | 2.399 (1.257–5.057), $p = 0.013$ | 2.071 (1.082–4.378), $p = 0.039$ |
| Only positive initial screening | 1.499 (0.825–3.062), $p = 0.221$ | 1.484 (0.816–3.033), $p = 0.234$ |
| *ICU admission* |  |  |
| NRS $\geq 3$ points | 2.144 (1.887–2.442), $p < 0.001$ | 1.850 (1.626–2.110), $p < 0.001$ |
| Only positive initial screening | 1.075 (0.957–1.212), $p = 0.229$ | 1.060 (0.942–1.196), $p = 0.340$ |

In Gamma regression with a log link, hospitalizations with NRS ≥3 points had a 1.66-fold expected LOS compared with the reference group (95% CI 1.57–1.76) after adjustment for age and sex, whereas hospitalizations with a positive initial screening only had a 0.83-fold expected LOS (95% CI 0.79–0.87).

## Discussion

In this large hospital-wide real-world cohort, three main findings emerged. First, elevated malnutrition risk identified by the NRS-2002 was associated with adverse clinical outcomes, including higher odds of in-hospital mortality, ICU admission, 30-day hospital readmission, and mechanical ventilation, as well as longer length of hospital stay. Second, a substantial number of hospitalizations had a positive initial screening but no documented final NRS-2002 assessment. This group showed higher frequencies of 30-day readmission, ICU admission, and in-hospital mortality than hospitalizations with a negative initial screening, although adjusted associations were not observed for all outcomes. Third, implementation of the intended two-stage screening pathway varied substantially across clinical departments. The comparison cohort from the pre-NRS period similarly showed higher frequencies of 30-day readmission and in-hospital mortality among hospitalizations with a positive versus negative initial screening. Together, these findings highlight both the clinical relevance of malnutrition risk identification and the challenges of implementing standardized screening in routine care.

In the main cohort, hospitalizations with NRS ≥3 points had substantially higher odds of in-hospital mortality, ICU admission, mechanical ventilation, and 30-day hospital readmission than those without elevated malnutrition risk, with associations remaining after adjustment for age and sex. They also showed a clinical profile characterized by older age, lower BMI, lower serum albumin concentrations, higher comorbidity burden, and a higher prevalence of malignant disease. These findings are consistent with previous evidence linking malnutrition risk and malnutrition to increased morbidity, mortality, and prolonged hospital stay [1, 17, 18, 19, 20, 21, 22]. The German Hospital Malnutrition Study similarly identified older age, malignant disease, and greater comorbidity as important determinants of hospital malnutrition and reported a 43% longer hospital stay among malnourished patients [3].

The subgroup of 21,842 hospitalizations with a positive initial screening but no documented final NRS-2002 assessment is particularly relevant to routine care. In the main cohort, this group showed higher frequencies of 30-day readmission, ICU admission, and in-hospital mortality than hospitalizations with a negative initial screening, while generally remaining below those observed among hospitalizations with NRS ≥3 points. Similar differences in 30-day readmission and in-hospital mortality were observed in the comparison cohort. However, without a completed final assessment, the underlying nutritional status of these hospitalizations cannot be determined. The substantial variation in final-assessment completion across discharge units therefore represents an important limitation of the screening pathway. Differences in implementation may affect both the identification of patients at malnutrition risk and the ascertainment of malnutrition risk across clinical settings. In addition, patient-reported information and changes in clinical status between the initial and final assessment may contribute to discrepancies between screening stages.

The extrapolated prevalence provides an estimate that accounts for hospitalizations with a positive initial screening but without a documented final assessment. It was calculated by applying the observed proportion of NRS ≥3 among completed assessments to hospitalizations without a documented final assessment. This estimate is therefore dependent on the assumption that assessed and unassessed hospitalizations have a similar distribution of malnutrition risk and should be interpreted accordingly. The distribution across discharge units further suggests that both clinical setting and screening implementation influence ascertainment, with the highest prevalence observed in oncology and palliative care. This is consistent with previous reports of increased nutritional vulnerability among patients with advanced or chronic diseases [21, 23, 24]. Among hospitalizations with a documented final NRS-2002 assessment, elevated malnutrition risk also increased with age and was highest among patients aged 70–79 years, consistent with previous reports [3, 21, 24, 25]. However, the NRS-2002 assigns an additional point to patients aged ≥70 years, meaning that part of this age-related pattern is inherent to the scoring system. As the analysis was performed at the hospitalization level, repeated admissions may additionally contribute to the observed association between age and malnutrition risk.

## Limitations

Several limitations should be considered. Incomplete and selective completion of the final NRS-2002 assessment may have introduced selection bias, and the extrapolated prevalence relies on the assumption that assessed and unassessed hospitalizations have a similar distribution of malnutrition risk. The retrospective single-center design, historical comparison cohort, and analysis at the hospitalization rather than patient level limit generalisability and may result in repeated representation of patients with frequent admissions. This may particularly affect the observed distributions of age and comorbidity burden, as patients with recurrent or complex hospitalizations may contribute multiple observations. In addition, models were adjusted for age and sex only, leaving potential residual confounding. Finally, nutritional interventions were not available, precluding assessment of their impact on clinical outcomes.

## Conclusion

This large hospital-wide real-world study found that elevated malnutrition risk identified by the NRS-2002 was associated with higher odds of in-hospital mortality, ICU admission, 30-day hospital readmission, and mechanical ventilation, as well as longer hospital stay. Hospitalizations with a positive initial screening but without a documented final assessment also showed higher frequencies of several adverse outcomes compared with those with a negative initial screening, highlighting the potential clinical relevance of the initial screening stage while underscoring the need for completion of the subsequent NRS-2002 assessment.

The findings further demonstrate substantial variation and incomplete implementation of the intended two-stage screening pathway across clinical departments. Consequently, observed NRS-based prevalence estimates are affected by selective assessment and should not be interpreted as unbiased estimates of hospital-wide malnutrition risk prevalence. More systematic and scalable approaches to malnutrition risk identification may help address gaps in routine screening, particularly in settings with limited resources. Data-driven approaches based on routinely collected clinical data could potentially complement existing screening workflows, but require external validation and prospective evaluation before implementation in clinical practice.

Future research should therefore address both reliable identification of patients at malnutrition risk and the implementation of standardized screening pathways, while linking risk identification to nutritional interventions and subsequent patient outcomes.

## Data Availability

The datasets analyzed during the current study are not publicly available because they contain sensitive patient-level health information and are subject to institutional and data protection restrictions. Aggregated data supporting the findings are reported in the manuscript and supplementary material.

## Competing Interests

The authors declare that they have no competing interests.

## Acknowledgements

None.

## CRediT authorship contribution statement

**Marie Gerwek:** Methodology, Data preprocessing, Outcome analysis, Writing – original draft, review and editing. **Silvia Walther:** Methodology, Data preprocessing, Descriptive analysis. **Ulrike Holdgrün:** Conceptualization, Data extraction, Data validation, Writing – review and editing. **Alicia Nierling:** Consultation. **Lars Selig:** Consultation. **Michael Stumvoll:** Consultation. **Toralf Kirsten:** Conceptualization, Supervision, Writing – review and editing. **Haiko Schlögl:** Conceptualization, Methodology, Consultation, Supervision, Writing – review and editing.

## Notes

### Competing Interest Statement

The authors have declared no competing interest.

### Author Declarations

Ethics Committee of University of Leipzig gave ethical approval for this work.

### Funding

- TMF – Technology and Methods Platform for Networked Medical Research e.V.

