## Supplementary Figures for "Nutritional Risk Screening and Clinical Outcomes in a Tertiary University Hospital: A Real-World Retrospective Cohort Study"

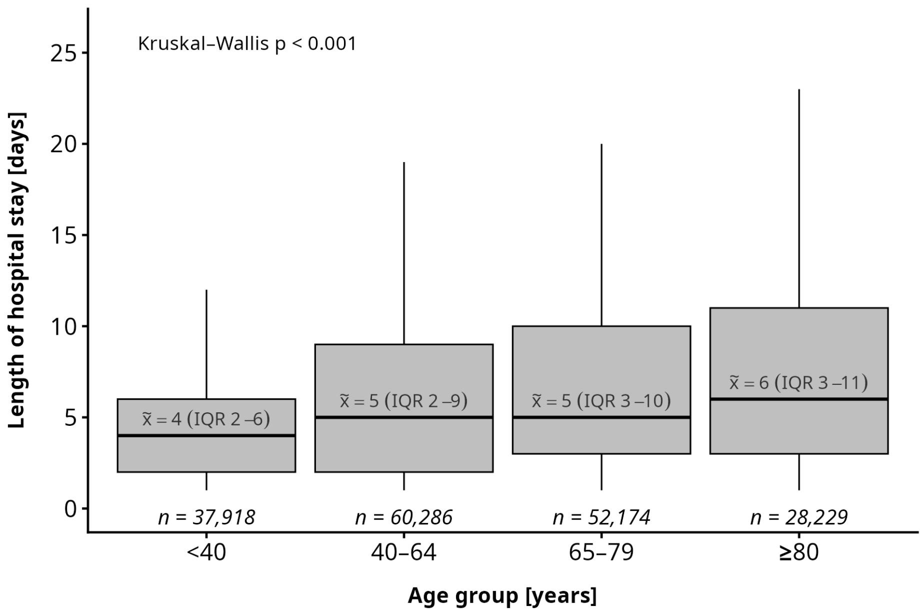


**Figure S1:** Length of hospital stay stratified by age group. Hospital length of stay was compared across four predefined age groups (<40, 40–64, 65–79, and ≥80 years) among all unique hospitalizations after the introduction of routine nutritional screening. Boxplots show the median, interquartile range (IQR), and whiskers extending to 1.5 × IQR, with outliers omitted for clarity. Median (IQR) and the number of hospitalizations are displayed for each age group. Statistical differences were evaluated using the Kruskal–Wallis test. IQR: interquartile range.

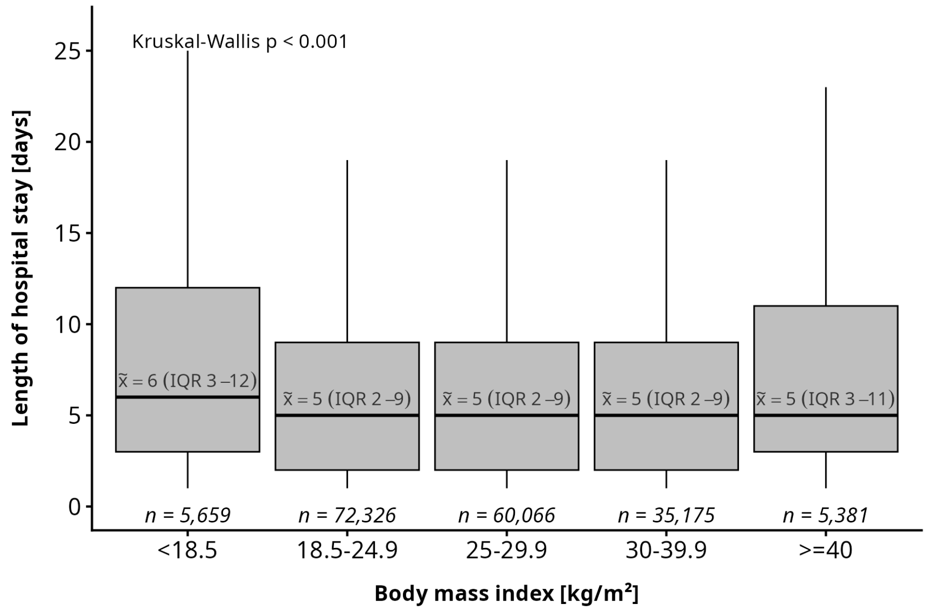


**Figure S2:** Length of hospital stay stratified by BMI category. Hospitalizations were categorized into five BMI groups (<18.5, 18.5–24.9, 25.0–29.9, 30.0–39.9, and ≥40.0 kg/$m^2$) among all unique hospitalizations after the introduction of routine nutritional screening. Boxplots show the median, interquartile range (IQR), and whiskers extending to 1.5 × IQR, with outliers omitted for clarity. Median (IQR) and the number of hospitalizations are displayed for each BMI group. Statistical differences were evaluated using the Kruskal–Wallis test. BMI: body mass index; IQR: interquartile range.

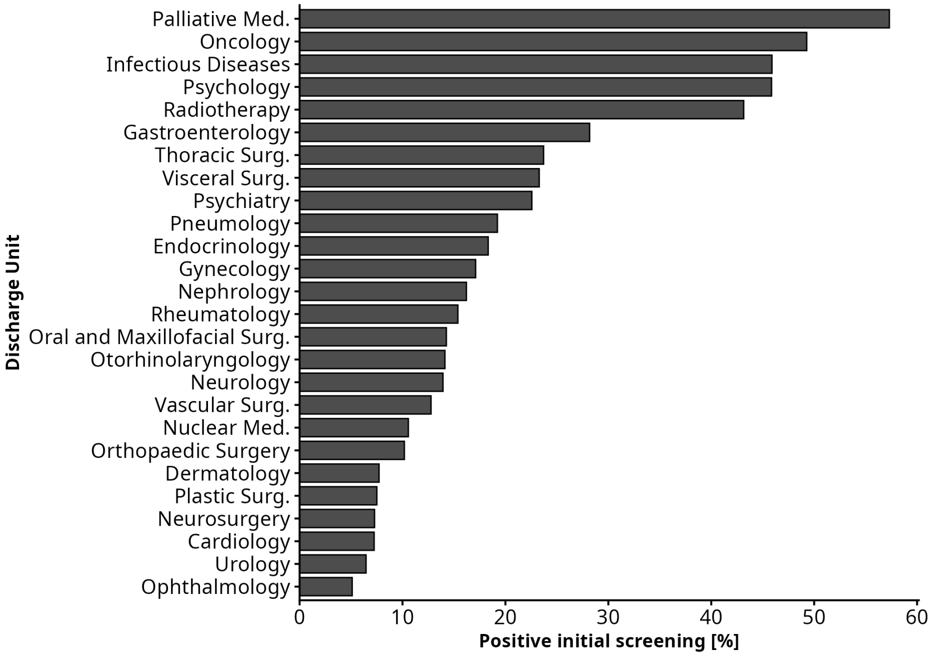


**Figure S3:** Proportion of hospitalizations with a positive initial nutritional screening across discharge units. The percentage of hospitalizations with a positive initial nutritional screening was calculated for each discharge unit among all unique hospitalizations after the introduction of routine nutritional screening. Only discharge units with at least 40 hospitalizations were included. Bars represent the proportion of hospitalizations with a positive initial screening, and percentages are displayed at the end of each bar.

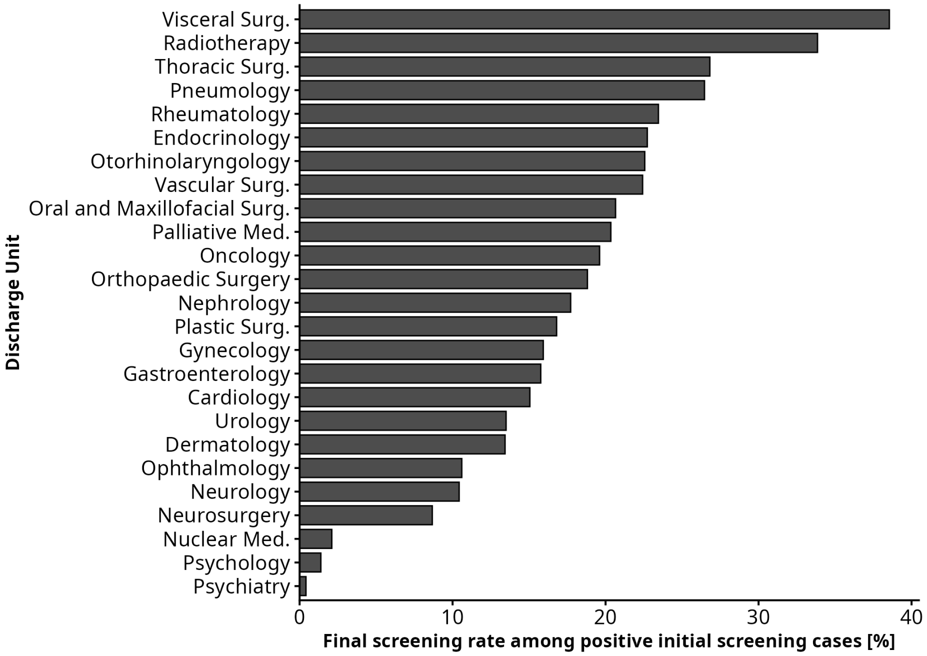


**Figure S4:** Proportion of hospitalizations with a positive initial screening and a completed final screening across discharge units. For each unit, the percentage of hospitalizations with an available NRS assessment was calculated among all hospitalizations with a positive initial screening result. Bars represent the proportion (%) of assessed hospitalizations within this subgroup. Only units with at least 40 hospitalizations with a positive initial screening were included. NRS: Nutritional Risk Screening 2002.

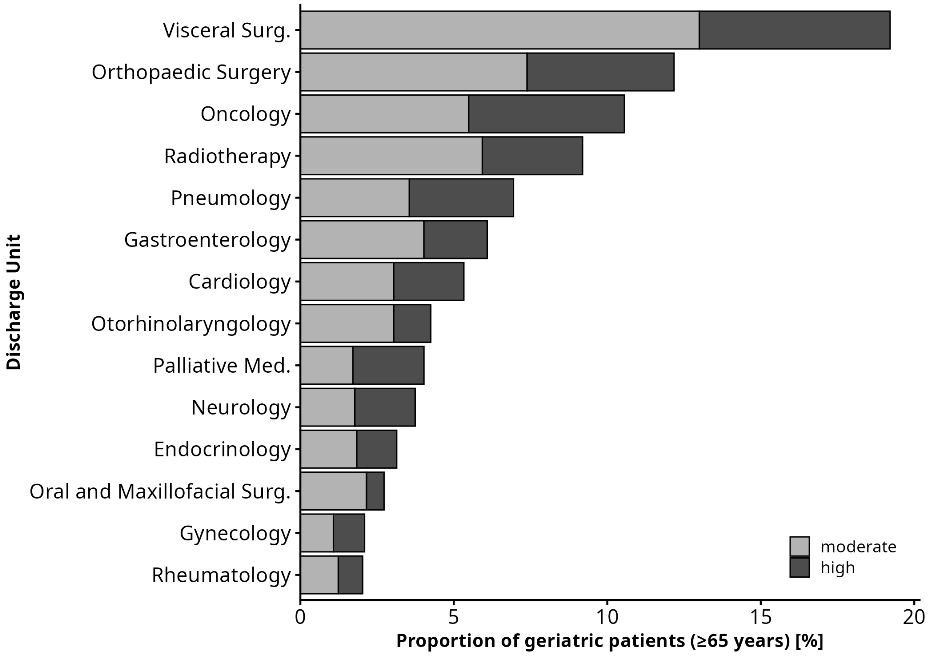


**Figure S5:** Hospitalizations among patients aged (≥65 years) with moderate (NRS 3–4 points) and high (NRS ≥5 points) malnutrition risk across discharge units. Percentages were calculated relative to all geriatric patients with documented final screening (NRS ≥3 points). Only discharge units with at least 50 hospitalizations were included.
